# The Seasonal End of Human Coronavirus Hospital Admissions with Implications for SARS-CoV-2

**DOI:** 10.1101/2020.05.15.20103416

**Authors:** Alan T. Evangelista

**Affiliations:** St. Christopher’s Hospital for Children, Department of Pathology and Laboratory Medicine, Philadelphia, PA, USA, Drexel University College of Medicine, Philadelphia, PA, USA

**Keywords:** Coronaviruses, Relative Humidity, SARS-CoV-2

## Abstract

The seasonality of influenza viruses and endemic human coronaviruses was tracked over an 8-year period to assess key epidemiologic reduction points in disease incidence for an urban area in the northeast United States. Patients admitted to a pediatric hospital with worsening respiratory symptoms were tested using a multiplex PCR assay from nasopharyngeal swabs. The additive seasonal effects of outdoor temperatures and indoor relative humidity (RH) were evaluated. The 8-year average peak activity of human coronaviruses occurred in the first week of January, when droplet and contact transmission was enabled by the low indoor RH of 20–30%. Previous studies have shown that an increase in RH to 50% has been associated with markedly reduced viability and transmission of influenza virus and animal coronaviruses. As disease incidence was reduced by 50% in early March, to 75% in early April, to greater than 99% at the end of April, a relationship was observed from colder temperatures in January with a low indoor RH to a gradual increase in outdoor temperatures in April with an indoor RH of 45–50%. As a lipid-bound, enveloped virus with similar size characteristics to endemic human coronaviruses, SARS-CoV-2 should be subject to the same dynamics of reduced viability and transmission with increased humidity. In addition to the major role of social distancing, the transition from lower to higher indoor RH with increasing outdoor temperatures could have an additive effect on the decrease in SARS-CoV-2 cases in May. Over the 8-year period of this study, human coronavirus activity was either zero or > 99% reduction in the months of June through September, and the implication would be that SARS-Cov-2 may follow a similar pattern.

## Introduction

The seasonality of respiratory viruses, such as influenza A and B and endemic human strains of coronaviruses (OC43, HKU1, NL63, 229E), is recognized in temperate regions of the globe, peaking in winter months. As stated by the WHO and CDC, these viruses as well as SARS-CoV-2 are primarily spread by droplet and contact transmission (1, 2). Studies of influenza viruses and animal coronaviruses have shown that droplet and contact transmission and virus viability is enhanced by cold temperatures, less than 5^0^C/41^0^F, and by low relative humidity (RH) of 20–30% (3, 4, 5). This lower humidity can be reached indoors during the winter months in northern temperate regions, January and February, due to the dryness of indoor heating. For example, when the cold outside temperatures in January and February range from –12^0^C/10^0^F to 5^0^C/41^0^F and the inside temperature is set to 22^0^C/72^0^F, the indoor RH can range from 20–30% and often can drop below 10% (6, 7). Transmission and infection with coronaviruses also is dependent on prolonged close contact between susceptible and infected individuals, touching contaminated surfaces, and on the immune status of the exposed host. It is well-recognized that social distancing, early case identification by testing, followed by isolation of index cases, contact tracing to identify and quarantine close contacts, and compliance with respiratory hygiene etiquettes are primary strategies for slowing the spread of respiratory viruses, including SARS-CoV-2. The objective of this study was to examine historical seasonal data from the past 8 years on the incidence of influenza and human coronavirus infections in pediatric patients requiring hospitalization. Key epidemiologic reduction points were recorded to determine the decline and seasonal end points of hospital admissions due to coronavirus infections. In addition, outdoor temperatures and indoor RH were monitored from January to May to determine the relationship of these environmental factors on the seasonal course of infection of human coronaviruses with implications for SARS-CoV-2.

## Materials and Methods

Data was analyzed for the 8-year seasonal influenza and human coronavirus period (Dec 2012-May 2020) at a pediatric hospital in north Philadelphia, PA, USA, a temperate zone region at 40^0^ North Latitude. A multiplex PCR (polymerase chain reaction) assay was used to detect 17 respiratory viruses and 3 bacteria (Respiratory Pathogens Panel, BioFire Diagnostics, Salt Lake City, UT) from nasopharyngeal swabs obtained from admitted patients with worsening respiratory symptoms. The percent weekly incidence of infection for influenza A, influenza B, and endemic human coronaviruses (HKU1, OC43, 229E, NL63) were determined from the number of admitted positive patients divided by the total number of admitted patients tested. Weekly disease incidence was recorded for each seasonal year and trending was assessed using 4 data points: week of peak activity, and weekly activity reduction points of 50%, 75%, and greater than 99%. Eight-year averages for the analysis points were determined as the mean weekly date and week of the month. The relationship of temperature and humidity on disease incidence was measured by recording weekly endemic coronavirus incidence for the 8-year period in comparison to weekly average outdoor temperature and relative humidity (RH) with a reference to measured indoor RH based on a study by Nguyen and colleagues (6). Weekly data points for each year were recorded as peak activity of infection and reduction points of 50%, 75%, and greater than 99%. Average weekly outdoor temperature and RH for Philadelphia were calculated from daily averages (8). Values of measured indoor RH obtained by Nguyen and colleagues (6) in the Boston area, 42^0^ N Latitude, were considered applicable to Philadelphia at 40^0^ N Latitude, both areas representing the metropolitan northeast temperate zone region of the United States. The use of aggregated patient viral incidence data in this study was reviewed and considered to be exempt from human subjects’ ethics review by the Institutional Review Board of Drexel University College of Medicine.

## Results

The 8-year averages for the week of peak viral activity were: first week in February for influenza A, last week in February for influenza B, and first week in January for the endemic strains of human coronaviruses (Table 1). The average decline for these viruses during the 8 seasons was determined at reduction points of 50%, 75%, and > 99%. For influenza A the 50%, 75% and > 99% average reduction points were the second week of March, fourth week of March, and the fourth week of April. For influenza B, there was a lag of 2 weeks for 50% and 75% reduction points to the fourth week of March, and the second week of April. The reduction point of > 99% for Influenza B also occurred in the fourth week of April. For the human coronaviruses the 50% and 75% reduction points were the first week of March and the first week of April, which occurred 12 weeks after the peak and was a slower course of decline than for Influenza A or B which occurred 7 weeks after peak activity. However, the > 99% average reduction point for human coronaviruses also occurred in the last week of April (Table 1). The seasonality of endemic coronaviruses activity was examined on a local level, by comparing the percent decline of infection to the outdoor temperature and outdoor RH in Philadelphia over the 8-year period, 2013–2020 (Table 2) and summarized with reference to indoor RH (Table 3). Outdoor RH fluctuated during the study period of January to April and no correlation was observed between viral activity and outdoor RH (Table 2). At average peak activity for coronaviruses in the first week of January, the average weekly outdoor temperature was 2^0^C/36^0^F and the accompanying indoor RH was 20–30% (Table 3) due to the drying effects of indoor heating (6, 7). At a 50% incidence reduction point in the first week of March, the average outdoor temperature was 7^0^C/45^0^F and the estimated indoor RH was 30–35% (6). At a 75% incidence reduction point in the second week of April the average outdoor temperature was 13^0^C/56^0^F with an estimated indoor RH of 35–45% (6). When the epidemiologic curve for coronaviruses decreased by greater than 99% in the last week of April, the average weekly outdoor temperature was 16^0^C/60^0^F and the indoor relative humidity had increased to 45–50% (6). Reductions of annual coronavirus activity, as measured by hospital admissions, correlated with increases in indoor RH from 30% to 50%. In Philadelphia, hospital admissions due to SARS-Co-V-2 peaked in mid-April 2020 and showed a steady decline into early May (9). The weekly human coronavirus incidence of hospital admissions was either zero or greater than a 99% reduction point at the end of April and into early May (Table 2) and was sustained from June to September in the 8–year study period (Table 3). Recurrence of human coronavirus activity was detected in October during the study period, as periods of indoor RH dropped below 50% (Table 3).

**Table 1.** Peak Incidence and Key Reduction Points of Influenza and Endemic Coronavirus Infections and Corresponding Week of Occurrence and Percent Incidence.

| Year | Incidence Status | Influenza A<br>Week of & % Incidence | Influenza B<br>Week of & % Incidence | Coronavirus<br>Week of & % Incidence |
| --- | --- | --- | --- | --- |
| 2020 | Peak | 2/16/20 – 18.5% | 12/22/19 – 19.3% | 1/26/20 – 5.0% |
|  | 50% Reduction | 3/08/20 – 10.4% | 2/02/20 – 10.3% | 2/09/20 – 2.0% |
|  | 75% Reduction | 3/22/20 – 0.8% | 3/08/20 – 4.6% | 3/08/20 – 1.2% |
|  | >99% Reduction | 4/05/20 – 0% | 4/05/20 – 0% | 4/12/20 – 0% |
| 2019 | Peak | 2/17/19 – 19% | 3/03/19 – 1.5% | 2/10/19 – 6.2% |
|  | 50% Reduction | 3/31/19 – 8.7% | 3/17/19 – 0.9% | 3/31/19 – 3.3% |
|  | 75% Reduction | 4/14/19 - 5.6% | 4/14/19 - 0% | 4/07/19 – 1.7% |
|  | >99% Reduction | 4/28/19 – 0% | 4/14/19 – 0% | 4/21/19 – 0% |
| 2018 | Peak | 2/04/18 – 10.8% | 2/04/18 – 9.0% | 12/10/17 – 15.8% |
|  | 50% Reduction | 3/11/18 – 6.0% | 4/15/18 – 4.2% | 1/07/18 – 7.8% |
|  | 75% Reduction | 4/15/18 – 2.1% | 4/22/18 – 3.3% | 1/28/18 – 5.2% |
|  | >99% Reduction | 4/22/18 – 0% | 4/29/18 - 0% | 3/18/18 - 0% |
| 2017 | Peak | 2/19/17 – 24.3% | 3/19/17 - 9.9% | 12/11/16 – 5.4% |
|  | 50% Reduction | 3/05/17 – 11.3% | 4/23/17 - 4.5% | 4/09/17 – 2.3% |
|  | 75% Reduction | 3/12/17 – 4.7% | 5/07/17 – 1.4% | 5/07/17 – 1.4% |
|  | >99% Reduction | 4/23/17 – 0% | 5/14/17 – 0% | 5/21/17 – 0% |
| 2016 | Peak | 3/06/16 – 20% | 4/03/16 – 5.8% | 1/31/16 - 7.1% |
|  | 50% Reduction | 3/27/16 – 10.1% | 4/17/16 – 3.8% | 3/20/16 - 2.9% |
|  | 75% Reduction | 4/10/16 – 5.0% | 5/01/16 – 1.1%- | 4/17/16 – 1.5% |
|  | >99% Reduction | 5/15/16 – 0% | 5/29/16 – 0% | 5/01/16 – 0% |
| 2015 | Peak | 1/11/15 – 12.8% | 2/22/15 -1.4% | 12/07/14 – 4.5% |
|  | 50% Reduction | 2/15/15 – 4.3% | 3/29/15 – 0.7% | 2/22/15 – 2.1% |
|  | 75% Reduction | 2/22/15 – 3.5% | 4/05/15 – 0% | 4/19/15 - 1.1% |
|  | >99% Reduction | 4/05/15 – 0% | 4/05/15 - 0% | 5/10/15 - 0% |
| 2014 | Peak | 1/26/14 – 33% | 3/30/14 - 6.6% | 1/19/14 – 9.6% |
|  | 50% Reduction | 4/06/14 - 9.8% | 4/27/14 – 3.6% | 3/16/14 - 5.7% |
|  | 75% Reduction | 4/20/14 – 5.7% | 5/04/14 – 1.8%% | 4/14/14 – 2.8% |
|  | >99% Reduction | 6/15/14 – 0% | 5/11/14 – 0% | 5/11/14 - 0% |
| 2013 | Peak | 1/06/13 - 22.6% | 2/10/13 – 16.6% | 1/20/13 – 4.7% |
|  | 50% Reduction | 1/27/13 – 7.3% | 3/10/13 – 6.8% | 2/27/13 – 2.4% |
|  | 75% Reduction | 2/03/13 – 5.2% | 3/31/13 – 3.8% | 4/13/13 – 1.9% |
|  | >99% Reduction | 3/10/13 – 0% | 4/21/13 – 0% | 4/21/13 – 0% |
|  | **8-Year Average** | **Influenza A** | **Influenza B** | **Coronaviruses** |
|  | Peak Activity | 2/05 (1<sup>st</sup> wk of Feb) | 2/24 (4<sup>th</sup> wk of Feb) | 1/09 (1<sup>st</sup> wk of Jan) |
|  | 50% Reduction | 3/09 (2<sup>nd</sup> wk of Mar) | 3/28 (4<sup>th</sup> wk of Mar) | 3/04 (1<sup>st</sup> wk of Mar) |
|  | 75% Reduction | 3/25 (4<sup>th</sup> wk of Mar) | 4/15 (2<sup>nd</sup> wk of Apr) | 4/14 (2<sup>nd</sup> wk of Apr) |
|  | >99% Reduction | 4/25 (4<sup>th</sup> wk of Apr) | 4/27 (4<sup>th</sup> wk of Apr) | 4/26 (4<sup>th</sup> wk of Apr) |

**Table 2.**
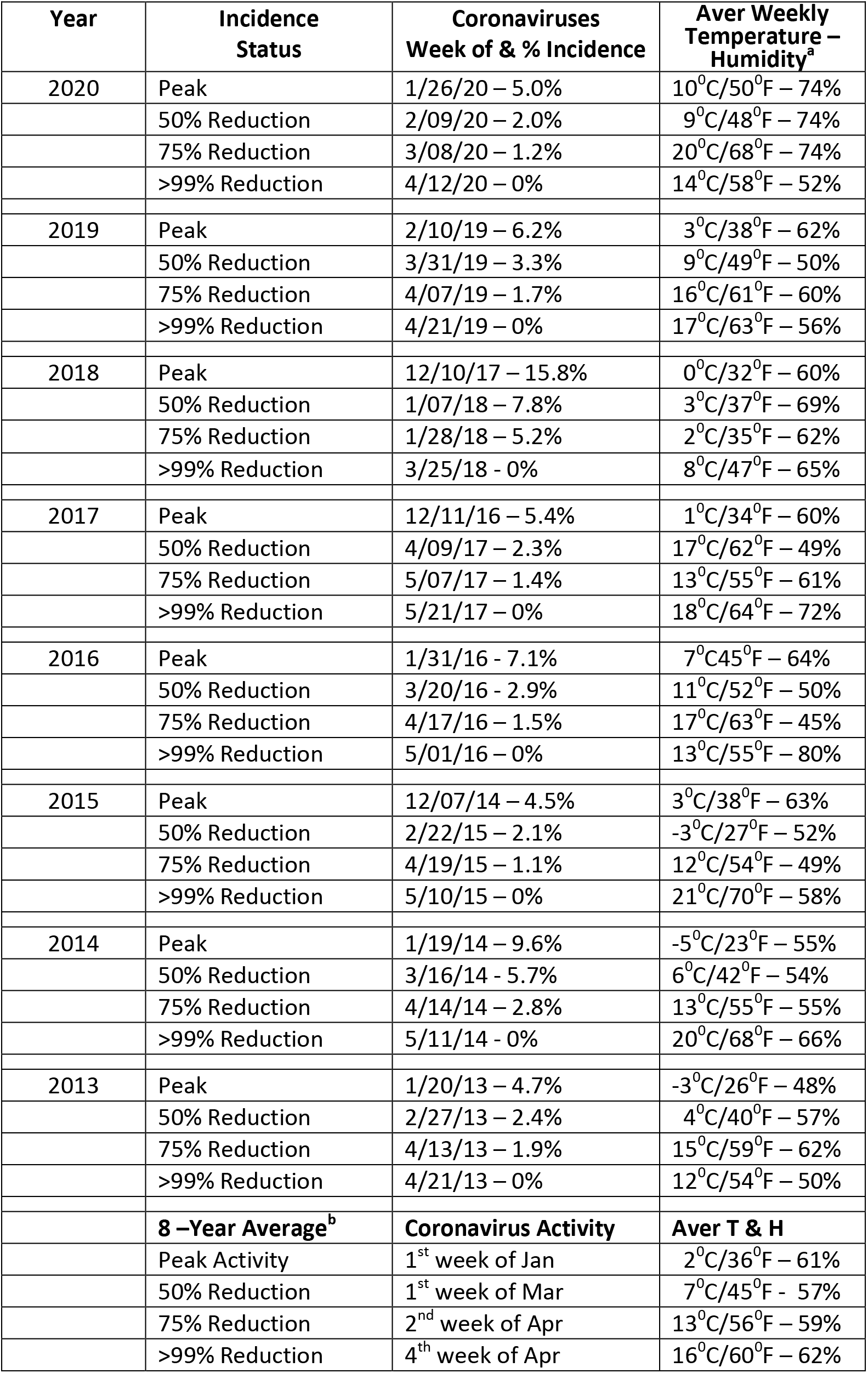
Percent Decline of Endemic Coronaviruses in Relation to Average.

**Table 3.** 8-Year Average Percent Activity of Endemic Coronaviruses in Relation to Average Weekly/Monthly Outdoor Temperature and Humidity and Estimated Indoor Relative Humidity.

| **Incidence Status** | **Coronaviruses**<br>**Average Week of**<br>**Activity** | **Aver Weekly**<br>**Outdoor**<br>**Temperature –**<br>**Humidity** | **Estimated**<br>**Indoor Relative**<br>**Humidity<sup>a</sup>** |
| --- | --- | --- | --- |
| Peak Activity | 1<sup>st</sup> week of Jan | 2<sup>0</sup>C/36<sup>0</sup>F – 61% | 20-30% |
| 50% Reduction | 1<sup>st</sup> week of Mar | 7<sup>0</sup>C/45<sup>0</sup>F - 57% | 30-35% |
| 75% Reduction | 2<sup>nd</sup> week of Apr | 13<sup>0</sup>C/56<sup>0</sup>F – 59% | 35-45% |
| >99% Reduction | 4<sup>th</sup> week of Apr | 16<sup>0</sup>C/60<sup>0</sup>F – 62% | 45-50% |
| Zero Activity or<br>>99% Reduction | Average for<br>month of May | 18<sup>0</sup>C/64<sup>0</sup>F – 63% | 45-55% |
| Zero or >99% Reduc | Aver for mo of June | 23<sup>0</sup>C/74<sup>0</sup>F – 64% | 45-60% |
| Zero or >99% Reduc | Aver for mo of July | 26<sup>0</sup>C/79<sup>0</sup>F – 64% | 50-60% |
| Zero or >99% Reduc | Aver for mo of Aug | 25<sup>0</sup>C/77<sup>0</sup>F – 66% | 50-65% |
| Zero or >99% Reduc | Aver for mo of Sept | 21<sup>0</sup>C/70<sup>0</sup>F – 68% | 50-65% |
| Initial Activity (1.3%) | Aver for mo of Oct | 14<sup>0</sup>C/58<sup>0</sup>F – 67% | 45-60% |
| Low Activity (2.4%) | Aver for mo of Nov | 9<sup>0</sup>C/48<sup>0</sup>F – 65% | 40-55% |

a. Estimate based on data from Nguyen, JL, et al.(6)

## Discussion

In temperate regions of the globe, the incidence of respiratory enveloped viruses, such as influenza and endemic human coronaviruses peak in winter months, usually in January and February, which is considered due to indoor social crowding with droplet and contact transmission and the added effect of low indoor relative humidity (RH). The decline of hospitalized patients admitted with human coronavirus infections, as recorded in Philadelphia, occurs in the months of March and April with an average decline of 75% occurring in early April and a decline of greater than 99% by the last week of April. One factor contributing to the percent decline of influenza viruses and coronaviruses is the transition from a low indoor RH (20–30%) in winter months with social crowding to an increase in indoor RH in March and April (35–50%) along with more outdoor exposure and the probable increase in social distancing. Monitoring changes in indoor RH from January through April is a key factor related to viral transmission, and it has been estimated that people in temperate zone metropolitan areas spend more than 90% of their time indoors (10). A study survey showed that 75% of homes and apartments in a temperate zone do not have humidifiers for winter months to maintain safe indoor RH at 30–50% (6). Indoor RH should not go above 60% because allergies and other respiratory problems may occur due to mold growth (7). In this study, increasing indoor RH from 30% to 50% corresponded with decreasing viral activity for both influenza and human coronaviruses (Table 3). Indoor RH is low and disparate compared to outdoor humidity in January and February, but becomes more aligned with outdoor humidity in April, May, and June with decreased use of indoor heating in northern temperate regions (6).

A RH of 50% has been shown to have a detrimental effect on the dynamics of droplet transmission and virus stability of influenza virus (3, 4). The in a study of animal coronaviruses, which were used as a model for SARS-CoV 2003, the greatest level of virus inactivation also was observed at 50% RH (5). When a person coughs, sneezes, or talks, the droplet size can range from 1–100 µm (11). Under drier conditions, such as 20–35% RH, the evaporation rate of the droplet occurs quickly, and the size of the droplet rapidly shrinks to below 10 µm (3, 4). At this small size the droplets can remain airborne for longer periods, increasing the time and distance over which transmission can occur. The rapid evaporation in dry air causes the NaCl concentrations within the droplet to crystallize out of solution quickly, maintaining virus stability (4). As the RH increases to 50%, the droplets evaporate more slowly and their larger size, 20–100 µm, causes them to settle out of the air rapidly. The slower evaporation process causes an increase in the concentration of NaCl within the droplet, resulting in increased osmotic pressure causing inactivation of an enveloped virus (3, 4). For example, in a study using influenza A virus spiked into mucus droplets, an increased RH from 48% to 52% resulted in a 10-fold (1 log) reduction in virus viability (4). When the RH was increased to 70% in the same study, an additional 1 log decrease in viability was noted (4). The relationship of influenza virus transmission and temperature/humidity also has been demonstrated using a guinea pig model showing that transmission proceeded more rapidly under cold, dry conditions (3). In contrast, under physiologic conditions as seen in the human respiratory tract with a RH of 99–100%, the NaCl concentrations remain at stationary levels and maintain the viability of influenza viruses. SARS CoV-2 causing COVID-19 is a new virus in humans, and has a higher estimated infectivity rate of R_0_ 2 – 2.5 compared to influenza virus of R_0_ 1.6 – 2 (12). However, the average size (125 nm) of SARS-CoV-2 and overall composition of this virus particle is similar to other coronaviruses (120–160 nm) examined in this study and the effects of humidity should apply to both types of enveloped (lipid-membrane bound) viruses.

The relationship of high seasonal winter activity of influenza and low humidity was demonstrated in a global study of temperate regions of the world with latitudes greater than 25^0^ N/S of the equator(13). The study also showed that when tropical countries within 10^0^ N/S latitude of the equator experienced their rainy season, their relative humidity was very high at 99–100%, and this corresponded to a higher influenza seasonal activity. These observations are consistent with a humidity range of 50–80% being detrimental to enveloped viruses and a very high humidity of 99–100% being protective of enveloped viruses, such as influenza. In additional, a temperature of 30^0^C/86^0^F has been reported to be detrimental to influenza virus, which is attained in summer months in northern hemisphere temperate regions (3).

Due to the sequential introduction of SARS-CoV-2 into different urban geographical locations in the United States in 2020, and the timing of physical distancing efforts, the epidemiologic curves have been different from the known seasonal coronaviruses. SARS-CoV-2 is a new virus and was believed to have been initially introduced into areas of the U.S. in Washington state, California, and New York City metropolitan areas in the February-March time frame, past the annual peak activity of endemic coronaviruses. However, the laws of physics regarding droplet transmission and viability should apply to similar viruses, and the increasing indoor RH to 50% could be a contributing factor to slowing down the transmission. It should be noted that outbreaks of COVID-19 occurred in March 2020 in southern cities with higher outdoor temperatures and humidity, such as Miami and New Orleans. These outbreaks most likely were due to close contact transmission (droplet and surface) that coincided with Spring Break and Mardi Gras with social crowding outdoors and indoors. As physical distancing continued through April and May, the number of hospital admissions for COVID-19 in these areas has decreased, as they have in Philadelphia (9).

The added effect of exposure to increasing outdoor temperatures, and the alignment of increasing indoor RH with outdoor RH going into the end of May and into June will continue to have a detrimental effect on coronaviruses viability.

Not only does increased humidity slow down respiratory viruses, it also has a beneficial effect on the innate immune response of the patient (3, 14). Since humans have not seen SARS-CoV-2 previously, they have no acquired immunity from memory T and B cells. Therefore, the innate immune response of mucociliary clearance is essential in preventing complications and the further progression of infection from the upper to the lower respiratory tract. As humidity and temperatures increase, mucus secretions increase in the nasopharynx, along with mucociliary clearance, phagocytosis of viruses by innate immune cells, and the activity of proteases against enveloped viruses (3). Mucus in the nasal passages facilitates the trapping of viruses and reduces the opportunity for viral adherence to target cells. One role of the ciliated epithelial cells in the nasopharynx is to remove viruses and bacteria, and these cells can be compromised by years of chemical damage, such as by smoking and exposure to air pollutants. Previous studies have shown that nasal mucociliary clearance is slower when breathing dry air (15, 16). The use of a humidifier, especially when sleeping at night, would add moisture to the air and help to improve mucociliary clearance. The ideal indoor relative humidity to protect health is considered to be between 30 and 50% (6). During the day, staying hydrated by periodically drinking water and warm liquids could help to increase nasal secretions. These actions could be helpful, since at the current time there are no approved antiviral therapies or vaccines, and maintaining a strong innate immunity is our primary defense. Other approaches such as practicing stress reduction techniques to boost immunity could be helpful, along with taking outside walks on warm humid days (while maintaining distancing) will have a dual effect of breathing fresh, more humid air, and having exposure to sunlight, which is antiviral and boosts natural levels of Vitamin D, helping the immune system.

In tracking human coronaviruses for the past 8 years, a seasonal pattern of decreased viral activity along with increasing indoor RH has been observed in the month of April in Philadelphia, PA, USA, a northeast city at 40^0^ North Latitude. The peak activity of SARS-CoV-2 occurred in early April 2020, in the northeastern US, which was three months later than endemic coronaviruses. However, the practice of physical distancing along has contributed to a reduction in hospital admissions of SARS-CoV-2 at the end of April and into May. Environmental factors that could have contributed to lower rates of infection could have been the increasing outdoor temperatures along with increasing indoor RH. In only three of eight years of tracking human coronaviruses did the greater than 99% reduction of activity continue through the middle of May, but in no years did it continue into June. Over the 8-year period, human coronavirus activity was either zero or > 99% reduction in the months of June through September, and the implication would be that SARS-CoV-2 would follow a similar pattern. There is always the possibility of local clusters of SARS-CoV-2 infections occurring as sporadic outbreaks throughout the summer. As with seasonal endemic coronaviruses, there is a reasonable possibility that SARS-CoV-2 will begin to circulate in northern temperate regions again in mid- to late October with the related cooler outdoor temperatures, use of indoor heating, and sporadic reduction in indoor RH below 50%.

A limitation of this study was that the percent incidence of seasonal coronavirus infection was based on pediatric hospital admissions. However, hospital admissions are representative of a smaller percentage of infections that are present in the community. As a novel virus, SARS-CoV-2 is more infectious than human coronaviruses, and may persist at very low levels in small local clusters from June through September, unrelated to environmental conditions.

## Conclusion

The major factor in reducing the epidemiologic curve for SARS-CoV-2, the viral agent of COVID-19, is social distancing. Other contributing factors in reducing the rates of transmission are adequate case testing, isolation, contact tracing, testing of contacts, and hand-washing. In addition, the patient’s innate immune response will play a major role in reducing the serious outcomes of infection. In addition to social distancing, the exposure to increased outdoor temperatures together with increased indoor RH should have an additive effect on decreasing the incidence of SARS-CoV-2 through May and into June. As a lipid-bound, enveloped virus, SARS-CoV-2 is similar in size to endemic coronaviruses and should be subject to similar dynamics of reduced droplet transmission and reduced virus viability as the indoor RH increases to 50%. Over the 8-year period of this study, human coronavirus activity was either zero or > 99% reduction in the months of June through September, and the implication would be that SARS-Cov-2 would follow a similar pattern.

## Data Availability

All data presented in 3 tables in manuscript. Original data sets available upon request.

## Ethics Statement

The use of aggregated patient viral incidence data in this study was reviewed and considered to be exempt from human subjects’ ethics review by the Institutional Review Board of Drexel University College of Medicine.

## Correspondence

Alan T. Evangelista, PhD, St. Christopher’s Hospital for Children, Department of Pathology and Laboratory Medicine – Microbiology & Virology, 160 East Erie Ave, Philadelphia, PA 19134, USA

## Acknowlegements

For consultation regarding this manuscript, I acknowledge Ishminder Kaur, MD (St. Christopher’s Hospital for Children, Section of Infectious Diseases). For their testing and surveillance efforts, I acknowledge Atif Abdalla, MS, Michael Heard, Julie Mathew, Susanne Dannert, MS, and Yolanda Innumerable (St. Christopher’s Hospital for Children, Department of Pathology & Laboratory Medicine – Virology). None received additional compensation beyond salary.

## ORCID

Alan Evangelista: http://orcid.org/0000-0001-5119-1167

